# Ultrasound to identify lupus patients with inflammatory joint symptoms with a better response to therapy: The USEFUL longitudinal multicentre study

**DOI:** 10.1101/2020.07.30.20142687

**Authors:** Khaled Mahmoud, Ahmed S Zayat, Md Yuzaiful Md Yusof, Katherine Dutton, Lee Suan Teh, Chee-Seng Yee, David D’Cruz, Nora Ng, David Isenberg, Coziana Ciurtin, Philip G Conaghan, Paul Emery, Christopher J Edwards, Elizabeth MA Hensor, Edward M Vital

**Affiliations:** NIHR Leeds Biomedical Research Centre, Leeds Teaching Hospitals NHS Trust, Leeds; Bradford Teaching Hospitals NHS Foundation Trust, Bradford; Royal Blackburn Teaching Hospital, Blackburn and University of Central Lancashire, Preston; Doncaster and Bassetlaw Teaching Hospitals NHS Foundation Trust, Doncaster; Guys and St Thomas Hospital, London; University College London, London; Centre for Adolescent Rheumatology, University College London, London; NIHR Clinical Research Facility, University Hospital Southampton NHS Foundation Trust, Southampton; Leeds Institute of Rheumatic and Musculoskeletal Medicine, University of Leeds, Leeds

**Author notes:** **Correspondence** Dr Edward Vital, Chapel Allerton Hospital, Leeds LS7 4SA, United Kingdom.

**Keywords:** Systemic Lupus Erythematosus, Ultrasound, Musculoskeletal Examination, Morning Stiffness, Corticosteroids

## Abstract

**Objective:** To determine whether SLE patients with inflammatory joint symptoms and ultrasound-synovitis achieve better clinical responses to glucocorticoid compared to patients with normal scans. Secondary objectives included identification of clinical features predicting ultrasound-synovitis.

**Methods:** A longitudinal muticentre study of SLE patients with physician-diagnosed inflammatory joint pain was undertaken. Clinical assessments, patient-reported outcomes, and bilateral hands and wrist ultrasound were collected at 0-, 2- and 6-weeks after intramuscular methylprednisolone 120mg. The primary outcome (determined via internal pilot analysis) was EMS-VAS at 2-weeks, adjusted for the baseline value, comparing patients with positive (GS≥2 and/or PD≥1) and negative ultrasound. Post-hoc analyses adjusting for fibromyalgia were performed.

**Results:** Of 133 patients recruited, 78/133 had positive ultrasound, but only 68% of these had ≥1 swollen joint. Of 66/133 patients with ≥1 swollen joint, 20% had negative ultrasound. Positive ultrasound was associated with joint swelling, symmetrical small joint distribution and serology. In full analysis set (n=133) there was no difference in baseline-adjusted EMS-VAS at week 2 (−7.7mm 95% CI − 19.0mm, 3.5mm, p=0.178). After excluding 32 fibromyalgia patients, response was significantly better in patients with positive ultrasound at baseline (baseline-adjusted EMS-VAS at 2-weeks - 12.1 mm, 95% CI −22.2mm, −0.1mm, p=0.049). This difference was greater when adjusted for treatment (−12.8mm (95% CI −22mm, −3mm), p=0.007). BILAG and SLEDAI responses were higher in ultrasound-positive patients.

**Conclusions:** In SLE patients without fibromyalgia, those with positive ultrasound had a better clinical response to therapy. Imaging-detected synovitis should be used to select SLE patients for therapy and enrich clinical trials.

## Introduction

Inflammatory joint disease affects more than 90% of patients with systemic lupus erythematosus (SLE) and is a major determinant of long term quality of life and disability[1, 2]. Our previous work suggests that this disproportionate impact on quality of life and disability may be due to under-treatment [1, 3]. SLE patients often have less joint swelling than rheumatoid or psoriatic arthritis patients.

In the British Isles Lupus Assessment Group (BILAG-2004) index[4], A and B scores (indicating active disease needing additional immunosuppression) can only be achieved if joints are swollen, w. The SELENA and 2K versions of the SLE Disease Activity Index (SLEDAI) score ‘arthritis’ if there are at least 3 or 2 inflamed joints, respectively[5, 6]. There is scope for interpretation of which signs qualify for this ‘inflamed’ criterion, but swelling is often sought before scoring this feature in clinical trials and routine practice. Patients who fail to score on these scales invariably fail to qualify for treatment with biologic therapies[7].

Previous ultrasound studies have shown that clinical joint swelling underestimates synovitis severity, although a systematic review found several methodological factors leading to uncertain estimates of the rates of ultrasound-synovitis[8]. We therefore conducted a large cross-sectional study to address these problems and estimated the rates of clinical and ultrasound abnormality in SLE patients presenting with inflammatory joint pain: 38% had clinical joint swelling, 35% had neither swelling on clinical examination nor evidence of ultrasound synovitis. However, 27% of patients had synovitis only detectable with ultrasound. Ultrasound-only synovitis appeared more clinically significant, being associated with worse tender joint count, physician VAS and serum IgG level[9].

We hypothesized that SLE patients presenting with inflammatory joint disease would demonstrate a better clinical response to therapy if synovitis was confirmed by ultrasound. To test this hypothesis we conducted a prospective, longitudinal, multi-centre study: UltraSound Evaluation For mUsculoskeletal Lupus (USEFUL). The primary objective was to determine whether patients with ultrasound-synovitis had better clinical responses to glucocorticoid therapy compared to patients without ultrasound synovitis. Other objectives included a comparison of ultrasound with the clinical variables at baseline to understand which are most useful when evaluating SLE patients.

## Methods

The study was approved by North West Greater Manchester Central Research Ethics Committee (Reference 16/NW/0060). Full details about the methodology can be found in online supplementary files.

### Patients and design

A prospective longitudinal UK multicentre cohort study was conducted in adults with SLE (meeting the revised ACR/SLICC 2012 criteria)[10], deemed by their physicians to have inflammatory joint pain requiring glucocorticoid therapy (swollen joints or specific BILAG-2004 / SLEDAI-2K scores were not required). Patients received intramuscular methylprednisolone 120mg at week 0 and had clinical and ultrasound assessments at weeks 0, 2 and 6 (Figure 1).

**Figure 1.**
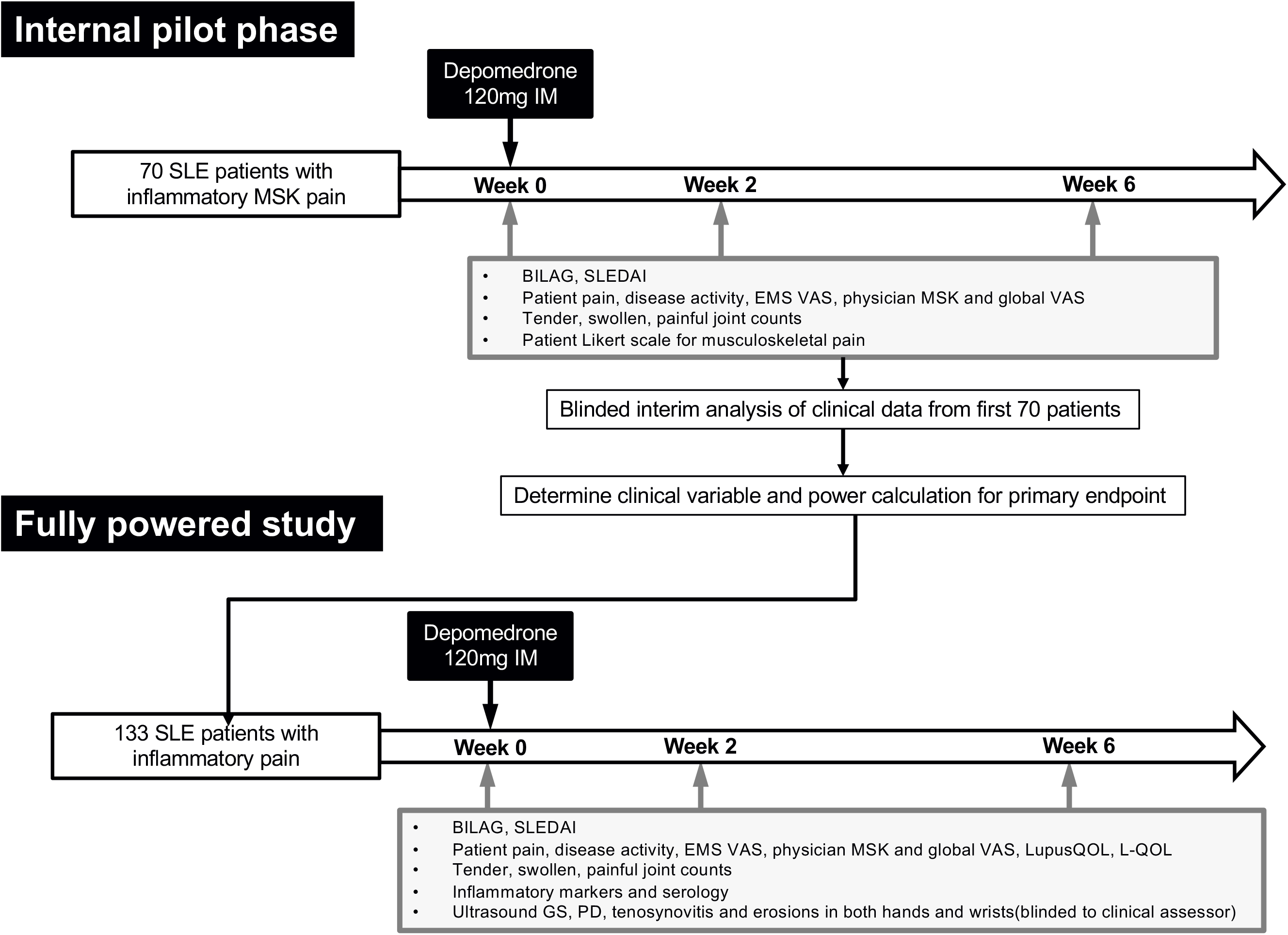
Study schematic. All patients followed the same treatment and assessment protocol. Clinical data shown from the first 70 patients was used to decide the primary clinical response variable and thereby calculate statistical power. Additional patients were then recruited to this target. Ultrasound data was not unblinded until all patients were recruited.

### Data and outcomes

Physician assessments included demographics, BILAG-2004, SLEDAI-2K, joint counts and global and musculoskeletal visual analogue scales (VAS), inflammatory markers and lupus serology and recorded features of inflammation, fibromyalgia, osteoarthritis, Early Morning Stiffness (EMS) and prior response to therapy. Patient-reported outcomes (LupusQoL, L-QoL, EMS minutes, EMS severity VAS, Likert scale for improvement in symptoms and Patient-acceptable symptom state) were collected[11]. Ultrasound was used to assess Grey scale (GS), Power Doppler (PD) and tenosynovitis according to OMERACT criteria[12] in both hands and wrists. Ultrasound was deemed “positive” if there was synovitis GS≥2 and/or PD≥1 or tenosynovitis GS≥1 and/or PD≥1. Physicians, ultrasonographers and patients were all blinded to each other’s assessments.

Details of ultrasound assessments are given in the supplementary methods. All sonographers attended a training event at baseline. The same four SLE patients were scored by each sonographer and proportions of agreement were calculated. These were Grey Scale: kappa = 0.69; Power Doppler: kappa = 0.98; Erosions: kappa = 0.85.

### Internal pilot

There were limited existing data on the most appropriate clinical measure of improvement[13]. We therefore conducted an internal pilot analysis of the first 70 patients (clinical data only). In this analysis, we evaluated various clinical variables with the best association with patient-reported improvement in symptoms based on Likert scale. This determined power calculations for the full study, and how many further patients should be recruited.

Change in the early morning stiffness severity based on VAS (EMS-VAS, mm) at week 2 from baseline (week 0) was selected as the primary outcome (see supplementary material). In other inflammatory arthritis this scale is better at discriminating high and low disease activity, and more responsive than EMS duration[14, 15]. We considered a difference of 20% compared to ultrasound-inactive to be the minimum difference of interest.

### Statistical Analyses

At baseline, descriptive data were presented on levels of agreement between ultrasound activity and joint swelling, BILAG-2004 and SLEDAI-2K musculoskeletal grades. In order to understand which symptoms, signs and routine laboratory tests were associated with ultrasound activity at baseline, we compared physician-reported features using Pearson’s Chi-squared for categorical variables or t-tests for continuous variables. These variables were also compared between SLEDAI-2K and BILAG-2004 categories.

The primary outcome was compared between patients with definite synovitis (ultrasound-active) and low level/no synovitis (ultrasound-inactive) at week 2 using quantile (median) regression, with cluster-robust standard errors employed to account for clustering of patients within centres. The primary analysis model adjusted for EMS-VAS at baseline; the unadjusted difference is presented for comparison.

In a planned sensitivity analysis, concomitant immunosuppressant and oral glucocorticoid (both recorded yes/no) were also added to the model. In a further planned sensitivity analysis, the above approaches were repeated in the per protocol set. Because there were a substantial number of patients with fibromyalgia, which may confound symptom responses, additional, unplanned, sensitivity analyses were performed in patients deemed not to have fibromyalgia at baseline[16].

The same analytical approach was then used to compare the other clinical variables at 2 and 6 weeks according to baseline ultrasound activity, controlling for baseline values of the outcome in each case. Because BILAG-2004, SLEDAI-2K and LupusQoL scores evaluate symptoms over the past month, we analysed these endpoints at week 6 only, using unadjusted changes from baseline between groups. An additional sensitivity analysis compared unadjusted changes from baseline between groups for all other clinical variables; this was added for comparison because baseline-adjusted analyses can potentially be biased when comparing nonrandomised groups.

## Results

133 SLE patients were recruited. 121 completed all visits (see supplement for details). Baseline characteristics are summarized in Table 1.

**Table 1:** Clinical characteristics and ultrasound findings at baseline

|  | All patients | US activity at baseline |  | P value |
| --- | --- | --- | --- | --- |
|  | N=133 | Inactive<br>N=55 | Active<br>N=78 |  |
| **Patient disposition** |  |  |  |  |
| Completed all study visits per protocol | 121/133 (91) | 52/55 (95) | 69/78 (88) | N/A |
| **Demographics** |  |  |  |  |
| Age (years)* | 46.1 (13.5) | 47.9 (12.3) | 44.8 (14.3) | 0.190 |
| Disease duration (years)* | 9.3 (8.9) | 10.2 (9.8) | 8.7 (8.1) | 0.352 |
| Male | 7/133 (5) | 0/55 (0) | 7/78 (9) | 0.022 |
| **Therapy** |  |  |  |  |
| NSAID or Cox-II inhibitor | 27/133 (20) | 12/55 (22) | 15/78 (19) | 0.421 |
| Prednisolone (maximum 5mg/day) | 31/133 (23) | 12/55 (22) | 19/78 (24) | 0.733 |
| Antimalarials | 89/133 (67) | 38/55 (69) | 51/78 (65) | 0.456 |
| Immunosuppressant (MMF, MTX, AZA) | 40/133 (30) | 17/55 (31) | 23/78 (29) | 0.125 |
| Biologic | 0 (0) | 0 (0) | 0 (0) | NA |
| **Inflammatory Features (physician rated)** |  |  |  |  |
| EMS | 115/133 (86) | 47/55 (85) | 68/78 (87) | 0.775 |
| Distribution | 113/133 (85) | 43/55 (78) | 70/78 (90) | 0.066 |
| Symmetry | 121/133 (91) | 46/55 (84) | 75/78 (96) | 0.013 |
| Swelling | 83/132 (63) | 28/54 (52) | 55/78 (71) | 0.029 |
| Serology | 87/130 (67) | 31/53 (58) | 56/77 (73) | 0.090 |
| Other lupus features | 66/133 (50) | 26/55 (47) | 40/78 (51) | 0.649 |
| Prior therapy response | 87/133 (65) | 39/55 (71) | 48/78 (62) | 0.263 |
| Jaccouds arthropathy | 6/133 (5) | 2/55 (4) | 4/78 (5) | 0.683 |
| Deformity | 6/133 (5) | 2/55 (4) | 4/78 (5) | 0.683 |
| Other lupus inflammatory | 3/133 (2) | 2/55 (4) | 1/78 (1) | 0.368 |
| **Fibromyalgia Features** |  |  |  |  |
| Overall opinion of fibromyalgia | 32/133 (24) | 14/55 (25) | 18/78 (23) | 0.752 |
| Fatigue | 30/132 (23) | 12/55 (22) | 18/77 (23) | 0.833 |
| Waking unrefreshed | 24/132 (18) | 9/55 (16) | 15/77 (19) | 0.647 |
| Cognitive symptoms | 20/132 (15) | 7/55 (13) | 13/77 (17) | 0.511 |
| Other somatic symptoms | 15/132 (11) | 8/55 (15) | 7/77 (9) | 0.330 |
| Associated disorders (e.g. IBS) | 5/133 (4) | 4/55 (7) | 1/78 (1) | 0.074 |
| **Osteoarthritis Features** |  |  |  |  |
| Overall opinion of osteoarthritis | 36/133 (27) | 16/55 (29) | 20/78 (26) | 0.659 |
| Hard tissue enlargement >1 joint | 25/133(19) | 13/54 (24) | 12/77 (16) | 0.224 |
| Hard tissue enlargement DIPs | 22/133(17) | 11/55 (20) | 11/77 (14) | 0.385 |
| Deformities consistent with OA | 14/133(11) | 6/55 (11) | 8/77 (10) | 0.924 |
| Previous radiographic evidence | 10/133(8) | 3/54 (6) | 7/74 (9) | 0.416 |
| Other OA features present | 5/133(4) | 1/54 (2) | 4/78 (5) | 0.332 |
| **Other musculoskeletal disorders** |  |  |  |  |
| Any other MSK disorder | 9/133 (7) | 4/55 | 5/78 | N/A |

2 All data are number of cases (%) and chi-squared tests except where marked \*, which indicates mean (SD) 3 and t-tests.

### Association of clinical features and ultrasound synovitis status

Active ultrasound was more likely if the clinical presentation included joint swelling, a symmetrical small joint distribution and active serology. Physician report of EMS, a common symptom used to identify inflammatory arthritis in routine practice, was not associated with ultrasound findings. There was also no association between active ultrasound findings and other SLE features, nor with the physician’s impression of their prior response to therapy (Table 1).

We found no association between ethnicity and ultrasound features, although low numbers of non-white non-south Asian patients limited this analysis. Although there were only 7 men in this study, all of them had ultrasound synovitis.

There was substantial disagreement between ultrasound and conventional clinical definitions of disease activity (Table 2).

**Table 2:** Agreement between clinical and ultrasound assessments at baseline

| A: Clinical joint swelling (any joint) versus ultrasound synovitis (hands/wrists) |  |  |  |  |
| --- | --- | --- | --- | --- |
| n (% of total) | ≥1 swollen joints | 0 swollen joints | Total |  |
| US inactive (<GS2 and <PD1 all joints) | 13 (10%) | 42 (32%) | 55 (41%) |  |
| US active (≥GS2 or ≥PD1 in ≥1 joint) | 53 (40%) | 25 (19%) | 78 (59%) |  |
| Total | 66 (50%) | 67 (50%) | 133 |  |
| B: Clinical joint swelling (hands/wrists) versus ultrasound synovitis (hands/wrists) |  |  |  |  |
| n (% of total) | ≥1 swollen joints | 0 swollen joints | Total |  |
| US inactive (<GS2 and <PD1 all joints) | 6 (4%) | 49 (37%) | 55 (41%) |  |
| US active (≥GS2 or ≥PD1 in ≥1 joint) | 54 (41%) | 24 (18%) | 78 (59%) |  |
| Total | 60 (45%) | 73 (55%) | 133 |  |
| C: SLEDAI-2K arthritis versus ultrasound synovitis |  |  |  |  |
| n (% of total) | Arthritis =Yes | Arthritis = No | Total |  |
| US inactive (<GS2 and <PD1 all joints) | 19 (14%) | 36 (27%) | 55 (41%) |  |
| US active (≥GS2 or ≥PD1 in ≥1 joint) | 59 (44%) | 19 (14%) | 78 (59%) |  |
| Total | 78 (59%) | 55 (41%) | 133 |  |
| D: BILAG-2004 musculoskeletal domain versus ultrasound synovitis |  |  |  |  |
| n (% of total) | BILAG A | BILAG B | BILAG C | Total |
| US inactive (<GS2 and <PD1 all joints) | 3 (2%) | 10 (7%) | 42 (32%) | 55 (41%) |
| US active (≥GS2 or ≥PD1 in ≥1 joint) | 11 (8%) | 42 (32%) | 25 (19%) | 78 (59%) |
| Total | 14 (11%) | 52 (39%) | 67 (50%) | 133 |

We compared swollen joint count in all joints with ultrasound scans in the hands and wrists (a typical ultrasound examination in routine practice, confirmed as clinically relevant in our previous study and systematic review {Zayat, 2018 #8;Zayat, 2016 #3}Table 2A). 13/66 (20%) of patients with clinical joint swelling did not have active ultrasound. 25/78 (32%) of patients with active ultrasound findings did not have joint swelling.

To evaluate the same set of joints, we evaluated clinical joint swelling in the 22 joints included in the ultrasound scan (Table 2B). This reduced the number of swollen joints not confirmed as having active ultrasound, but clinical examination remained insensitive.

19/78 (24%) of patients scored for arthritis on the SLEDAI-2K did not have active ultrasound. Equally, 19/78 (24%) of patients with active ultrasound were not scored for arthritis on the SLEDAI-2K.

13/66 (20%) of patients who scored A or B for the musculoskeletal BILAG-2004 did not have active ultrasound. 25/78 (32%) patients with active ultrasound did not score A or B on the musculoskeletal domain of the BILAG-2004. BILAG-2004 includes a “C” grade representing inflammatory pain. All patients in the study met this grade since it matched the eligibility criteria. However, notably, the majority of patients with BILAG-20004 C (42/67, 63%) did not actually have active ultrasound findings. It must be noted that BILAG-2004 and SLEDAI-2K scores could also be influenced by joints that were not included in the ultrasound examination.

Patients with active ultrasound had statistically significant worse symptoms and signs of lupus arthritis (Table S2), and higher ESR and total IgG level (Table S4). Active ultrasound was also associated with anti-Sm and anti-RNP antibodies (Table S4). 20/26 (77%) of Sm+ patients had active ultrasound, compared to 58/107 (55%) of Sm-patients (p = 0.035). Similarly, 17/22 (77%) of RNP+ patients had active ultrasound compared to 61/111 (55%) of RNP-patients (p = 0.052).

Quality of life scores as measured by the LupusQoL was similar in patients with active and inactive ultrasound (Table S3). Exclusion of patients with fibromyalgia did not change this result (data not shown). All patients in this study had active joint symptoms requiring increased therapy at baseline, therefore they would be expected to rate their quality of life as being affected even if this was not due to active inflammation.Primary outcome

Results for primary outcome analysis is shown in Figure 2 with full statistical data in Table S7.

**Figure 2.**
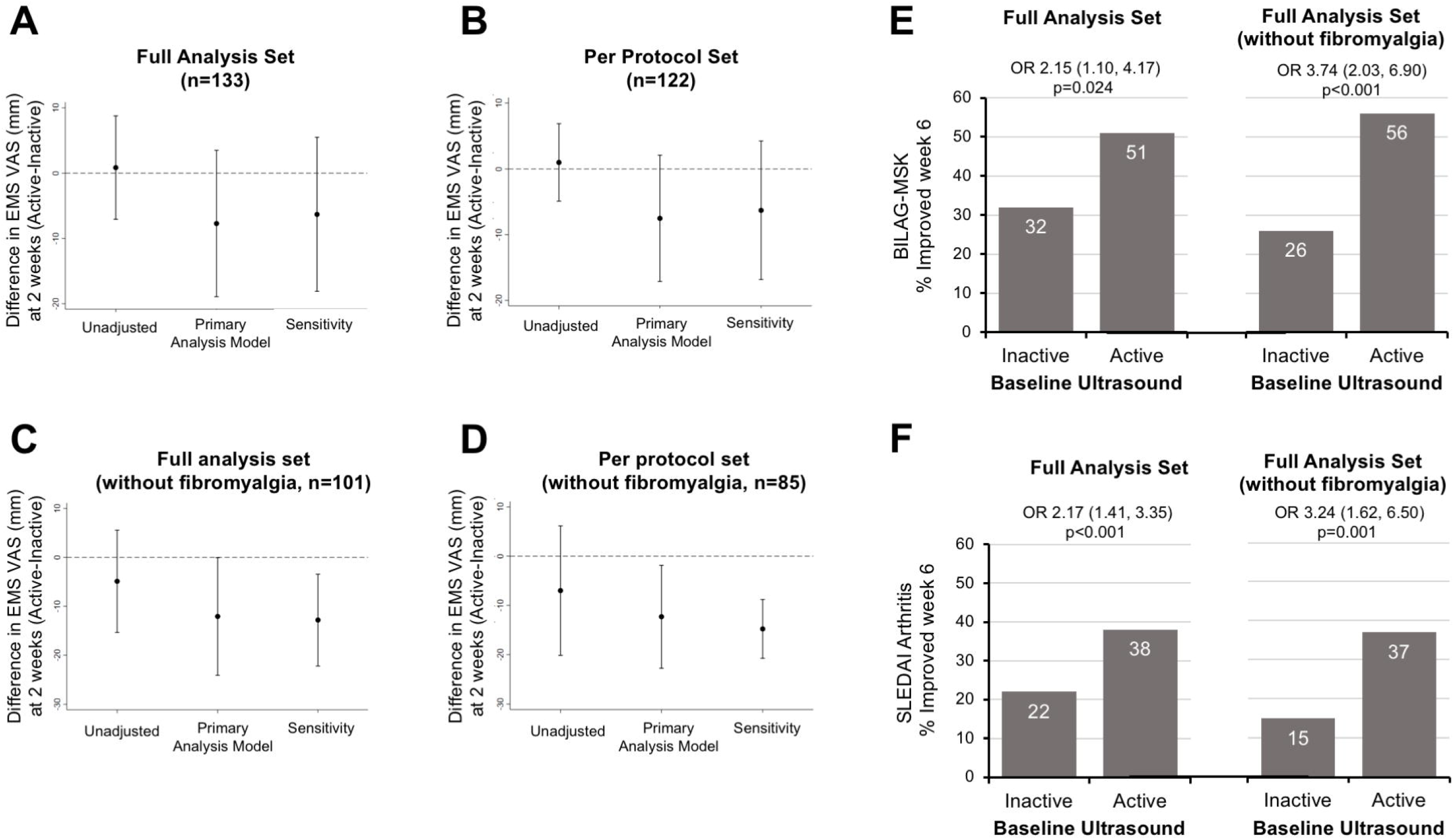
Primary endpoint; clinical response according to baseline ultrasound. A-D show the primary efficacy variable (Early Morning Stiffness VAS at week 2) according to baseline ultrasound status. Horizontal dotted line indicates no difference between ultrasound active and inactive groups. Error bars below this dotted line indicate a better response in ultrasound active patients. Unadjusted values do not account for baseline differences in EMS VAS between ultrasound active and inactive patients. The primary analysis model was adjusted for this baseline difference. The sensitivity analysis was also adjusted for use of NSAIDs, prednisolone and immunosuppressants. E-H show improvement in MSK components of BILAG and SLEDAI according to baseline ultrasound status. Parts E shows percentage of patients with improvement in the musculoskeletal component of the BILAG. Improvement was defined as reduction by at least one grade (i.e. A to B, B to C, or C to D). Part F shows the percentage of patients with improvement in the musculoskeletal items on the SLEDAI (arthritis and myositis, although no patient in this study was scored for myositis). Improvement was therefore defined as resolution of arthritis (reduction from 4 points to 0 points).

In the full analysis set (n=133) the mean (SD) baseline EMS-VAS was 57.7mm (4.1) in ultrasound-inactive patients, and 58.6mm (5.9) in ultrasound-active patients. The primary efficacy analysis showed no clinically or statistically significant difference between these groups (baseline-adjusted difference −7.7mm (95% CI −19.0, 3.5), p=0.178). The planned sensitivity analysis, which was also adjusted for immunosuppressant and oral glucocorticoid use, did not substantially affect this result (−6.3mm (95% CI −18.1, 5.5), p=0.293). Results in the per-protocol set (n=122) were broadly similar.

### Post-hoc analyses in patients without concurrent fibromyalgia

32 patients had a clinician diagnosis of fibromyalgia at baseline (which generally does not respond to glucocorticoids[17]). In patients without fibromyalgia (n=101), mean baseline EMS-VAS was 56.3mm (4.5) in ultrasound-inactive patients and 51.4mm (6.0) in ultrasound-active patients. An unplanned sensitivity analysis, repeating the baseline-adjusted primary analysis in this group, showed a significantly lower EMS-VAS at 2 weeks in patients with ultrasound synovitis at baseline (ultrasound-active – ultrasound-inactive −12.1 (95% CI −24.1, −0.1), p=0.049). This difference was greater in the treatment-adjusted sensitivity analysis (−12.8 (95% CI −22.2, −3.44), p=0.007) and in the per-protocol treatment-adjusted sensitivity analysis (−14.8 (95% CI −20.8, −8.8), p<0.001).

### Other clinical endpoints

BILAG-2004 and SLEDAI indices (which include manifestations from the previous 30 days) at 6 weeks according to baseline ultrasound activity are shown in Figure 3 and Tables S12-13. In the full analysis set (n=133) the musculoskeletal-BILAG improved (reduced by at least one grade, i.e. A to B, B to C or C to D) in 32% of ultrasound-inactive patients and 51% of ultrasound-active patients (odds ratio (OR) 2.15 (1.10, 4.17), p=0.024). The SLEDAI-2K arthritis criterion improved (resolution of arthritis criterion) in 22% of ultrasound-inactive patients and 38% of ultrasound-active patients (OR 2.17 (95% CI 1.41, 3.35), p<0.001).

**Figure 3.**
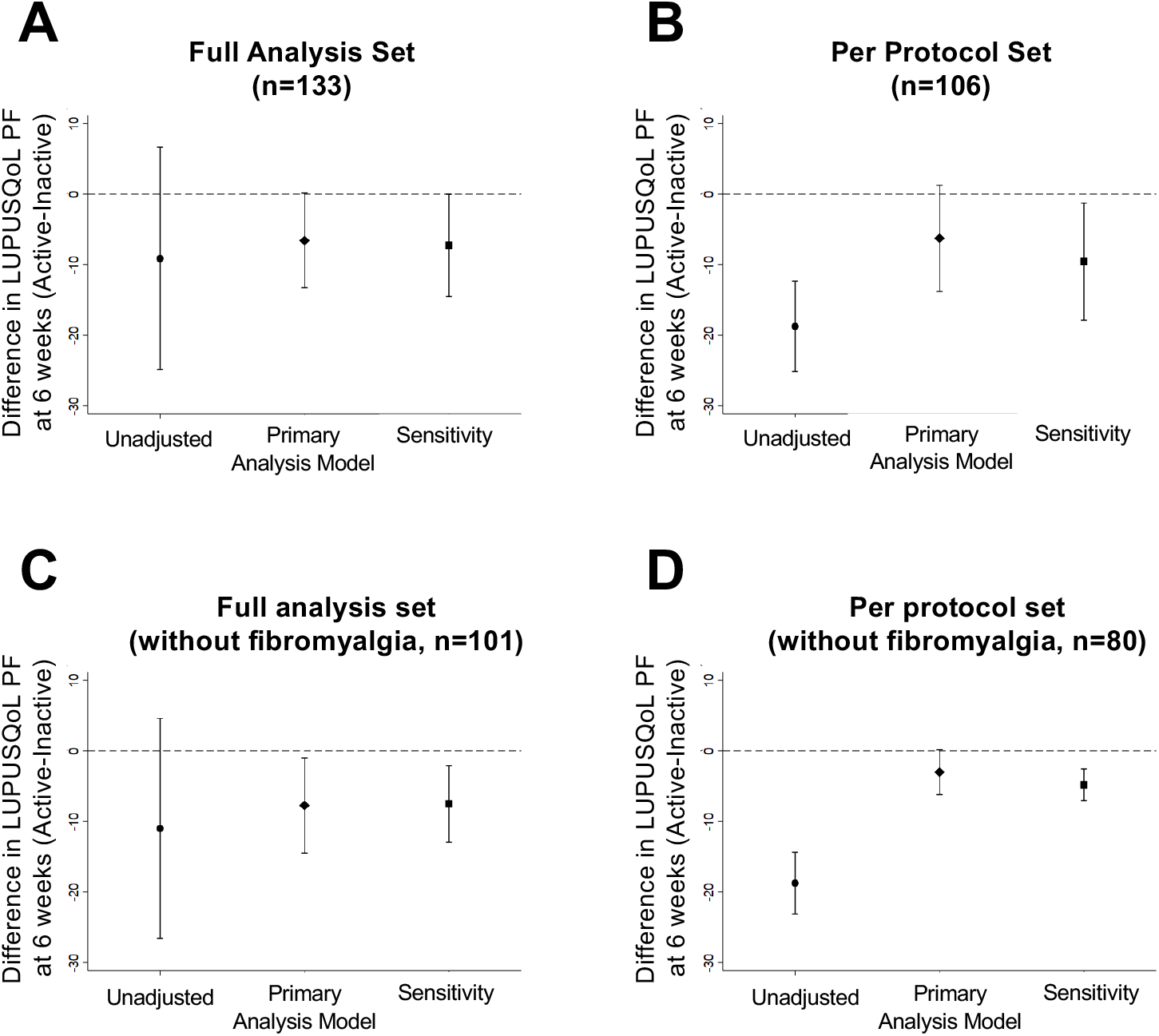
Change in LupusQoL physical function domain according to baseline ultrasound status. Horizontal dotted line indicates no difference between ultrasound active and inactive groups. Error bars below this dotted line indicate a better response in ultrasound active patients. Unadjusted values do not account for baseline differences in the physical function score between ultrasound active and inactive patients. A. The primary analysis model was adjusted for this baseline difference. The sensitivity analysis was also adjusted for use of NSAIDs, prednisolone and immunosuppressants. B shows the same analysis for the per-protocol set. C shows the same analysis for the study population excluding patients with fibromyalgia. D shows the same analysis for the per protocol set excluding fibromyalgia.

The improved responses in ultrasound-active patients were again more evident in patients without fibromyalgia (n=101). The MSK-BILAG-2004 improved in 26% of ultrasound-inactive patients and 56% of ultrasound-active (OR 3.74 (95% CI 2.03, 6.90), p<0.001). The MSK-SLEDAI-2K improved in 15% of ultrasound-inactive patients and 37% of ultrasound-active patients (3.24 (95% CI 1.62, 6.50), p=0.001).

Data for other clinical outcomes at 2 and 6 weeks is shown in supplementary tables S8 and S9.

### Quality of life

Three domains of the LupusQoL also showed greater improvement in ultrasound-active patients (Table 3 and Figure 3). Significantly greater improvement was seen in ultrasound-active patients for the physical health, burden to others, and body image domains. The size of these effects differed slightly between the 2 and 6 week timepoints, and between the primary and sensitivity analysis. However, differences were found in both the full analysis set and the subgroup without fibromyalgia. Patients without fibromyalgia were significantly more likely to achieve a patient-acceptable symptom state at week 2 if ultrasound was active at baseline (38% versus 32%, baseline-adjusted primary OR 1.75 (1.22, 2.49), p=0.002).

**Table 3:** LupusQoL improvements according to baseline ultrasound status

| Variable | 2 weeks |  |  |  | 6 weeks |  |  |  |
| --- | --- | --- | --- | --- | --- | --- | --- | --- |
|  | Adjusted primary |  | Adjusted sensitivity |  | Adjusted primary |  | Adjusted sensitivity |  |
|  | Difference (95% CI) | p | Difference (95% CI) | p | Difference (95% CI) | p | Difference (95% CI) | p |
| **All patients (n=133)** |  |  |  |  |  |  |  |  |
| Physical health | -0.45 (-3.93, 3.03) | 0.799 | -0.57 (-4.10, 2.95) | 0.749 | -6.57 (-13.27, 0.13) | 0.054 | -7.25 (-14.48, -0.01) | 0.050 |
| Pain | 0.65 (-3.84, 5.15) | 0.776 | 1.28 (-3.74, 6.30) | 0.616 | -5.22 (-13.60, 3.16) | 0.218 | -3.07 (-10.18, 4.04) | 0.395 |
| Planning | -0.17 (-5.35, 5.00) | 0.948 | -0.14 (-4.58, 4.31) | 0.951 | -1.75 (-9.93, 6.43) | 0.673 | -2.22 (-9.63, 5.20) | 0.554 |
| Burden to others | -8.33 (-18.45, 1.78) | 0.106 | -8.33 (-16.68, 0.01) | 0.050 | -7.54 (-15.27, 0.18) | 0.056 | -5.67 (-11.68, 0.34) | 0.064 |
| Emotional health | 0.05 (-3.30, 3.39) | 0.978 | 0.18 (-3.33, 3.68) | 0.922 | -3.61 (-9.09, 1.88) | 0.195 | -2.60 (-8.44, 3.24) | 0.379 |
| Body image | -0.51 (-6.57, 5.54) | 0.867 | -0.71 (-7.01, 5.58) | 0.823 | -8.83 (-16.65, -1.01) | 0.027 | -7.95 (-15.64, -0.26) | 0.043 |
| Fatigue | 1.42 (-3.41, 6.26) | 0.562 | 0.86 (-3.04, 4.75) | 0.665 | -2.56 (-8.15, 3.03) | 0.366 | -2.78 (-7.95, 2.40) | 0.291 |
| **No fibromyalgia (n=101)** |  |  |  |  |  |  |  |  |
| Physical health | -1.37 (-5.58, 2.85) | 0.514 | -0.93 (-5.33, 3.47) | 0.671 | -7.74 (-14.47, -1.00) | 0.025 | -7.49 (-12.90, -2.07) | 0.008 |
| Pain | 5.24 (-2.32, 12.79) | 0.167 | 3.61 (-2.22, 9.43) | 0.221 | -7.09 (-16.12, 1.93) | 0.121 | -4.49 (-14.43, 5.44) | 0.364 |
| Planning | -8.33 (-13.57, -3.09) | 0.002 | -7.85 (-12.96, -0.75) | 0.003 | -1.84 (-10.70, 7.02) | 0.682 | -1.71 (-9.61, 6.19) | 0.670 |
| Burden to others | 1.19 (-4.33, 6.70) | 0.671 | 1.57 (-3.64, 6.78) | 0.551 | -8.36 (-16.05, -0.67) | 0.033 | -6.68 (-13.28, -0.08) | 0.047 |
| Emotional health | 0.69 (-6.38, 7.75) | 0.846 | -1.10 (-10.00, 7.81) | 0.805 | -2.96 (-9.39, 3.47) | 0.366 | -0.41 (-6.38, 5.56) | 0.893 |
| Body image | 1.89 (-3.49, 7.27) | 0.486 | 1.25 (-3.61, 6.10) | 0.613 | -9.02 (-20.08, 2.03) | 0.108 | -7.72 (-16.46, 1.02) | 0.083 |
| Fatigue | -0.70 (-2.28, 0.89) | 0.386 | -0.50 (-1.72, 0.72) | 0.414 | -4.29 (-12.27, 3.68) | 0.289 | -3.96 (-12.57, 4.64) | 0.364 |

3 Adjusted primary analysis is difference between medians of ultrasound-active and ultrasound-inactive patients, adjusted for baseline value of the outcome variable. 4 Adjusted sensitivity analysis is also adjusted for immunosuppressant and oral glucocorticoid use. Negative values indicate a greater improvement in patients with 5 active ultrasound at baseline. Due to failure of the analysis models to converge, possibly due to the large number of patients who said the domain was not 6 applicable, data has not been provided for the LupusQoL domain of Intimate relationship

### Sensitivity analyses for secondary clinical endpoints

Sensitivity analyses comparing unadjusted changes from baseline between groups (Tables S10-S13) generally indicated the same direction of effect for primary and secondary outcomes compared to the main analysis. When patients with fibromyalgia were excluded, both analysis methods agreed the response was greater in those with baseline ultrasound-active for EMS VAS and the LupusQoL domain ‘burden to others’ as well as SLEDAI MSK at week 6. However, for physician global and MSK VAS the two analysis methods disagreed over the direction of effect.

## Discussion

This study the most comprehensive clinical and imaging data to define which SLE patients with inflammatory joint symptoms are most responsive to glucocorticoid therapy. We demonstrated disagreement between ultrasound synovitis and features commonly used to guide immunosuppressive therapy: joint swelling or completion of the BILAG-2004 and SLEDAI-2K indices. Although the primary outcome was not met, this may be due to a large proportion of patients having fibromyalgia. When they were excluded, we found a greater clinical response to glucocorticoid in participants with ultrasound synovitis at baseline in several clinical and patient-reported endpoints. These results have importance for the selection of patients for therapy in routine practice, clinical trials in SLE, and for the utility of ultrasound in this disease.

Despite the availability of licensed immunosuppressive therapies, musculoskeletal symptoms continue to have a major negative impact on quality of life in patients with SLE[1]. Our results indicate that outcomes may improve by targeting treatment to appropriate patients. In patients with no swelling but positive ultrasound synovitis, there may be value in escalating immunosuppressive therapy. Conversely, a negative ultrasound may indicate that it is safe to continue to taper glucocorticoids, which is important given the toxicity of long-term glucocorticoids.

If ultrasound is not available, the most useful signs indicating active joint inflammation are distribution, swelling and symmetry. Conversely, some other symptoms (EMS, other lupus features, prior therapy response) commonly used in practice may not be helpful. Consistent with our previous work, the level of IgG may be useful in identifing active lupus arthritis[9]. Sm and RNP antibodies are also helpful.

Despite the negative impact of fibromyalgia on responsiveness, it is important to note that we confirmed ultrasound-proven inflammation in patients who also had this condition, as well as osteoarthritis or other non-inflammatory pathologies. There may be longer term benefits of treatment in these subgroups and the value of treating ultrasound-synovitis in patients with fibromyalgia requires further work. The number of patients diagnosed with fibromyalgia in our study was high compared to unselected SLE cohorts[18], which is not suprising since the main inclusion criterion was pain.

The data from this study fulfill many of the criteria in the OMERACT filter for validation of ultrasound as an outcome measure in SLE[19]. Previous data had shown that ultrasound had face validity, and concurrent validity (association with BILAG-2004 and SLEDAI indices [8, 20, 21]. In the present study, for the first time we demonstrate that it has predictive validity for therapy response.

Many clinical trials of new therapies in SLE failed to demonstrate any benefit even though there were reasons to believe these therapies should have been effective[22]. In addition phase II and phase III trial data using the same monoclonal have produced very different results. Thus the primary endpoint was not met in the phase II trials of belimumab, but was met in phase III trials that altered eligibility criteria and the primary endpoint[23-25]. A trial of rituximab in non-renal SLE was negative, but the therapy is widely used in routine practice based on extensive open label evidence of efficacy[26][27-29]. One phase III trial of anifrolumab was negative, despite strongly positive phase II data and another phase III trial was positive [30, 31] For belimumab, markers of greater disease activity at baseline predicted a larger effect size between active and placebo groups. Our results help to define a target population to enroll or enrich clinical trials with confirmed synovitis and consequent greater response to therapy. Usually eligibility is based on clinical examination for swollen joints. We showed that 20% of such patients do not have active synovitis. Further, we showed that, although patients with fibromyalgia may have confirmed active synovitis, this comorbidity confounds assessment of response. Hence our results suggest that the use of ultrasound (or other imaging) synovitis as an inclusion criterion, and exclusion of patients with significant fibromyalgia that would affect disease activity assessment may result in larger effect sizes in clinical trials of new therapies.

The main limitation of our study is the open label design. This limits understanding of how these patient selection criteria would function in an RCT. Although ultrasound is a more sensitive test of inflammation than clinical examination, there could be immune-mediated causes of pain that are not well captured by this tool, such as bone oedema[32]. The fibromyalgia analysis was post-hoc. However, it is logical in line with clinical practice since fibromyalgia is a well-known co-morbidity that influences response to therapy[33]. Lastly, a possible explanation for the discrepancy between ultrasound and BILAG-2004 and SLEDAI was that ultrasound was only done on hands while the BILAG and SLEDAI indices take into account all joints. BILAG-2004 and SLEDAI are also dependent on the training of the assessor. Our study was conducted in BILAG member centres. In a less specialist setting, these instruments may have performed worse compared to ultrasound.

Future research should evaluate the use of ultrasound in randomised trials to determine whether it can measure and stratify differences between treatment groups. Such an RCT is in progress (ROOTS, NCT03054259). Future work will define clinical outcome measures for musculoskeletal disease activity that better match the results obtained from ultrasound.

In conclusion, ultrasound-synovitis is clinically important and responsive to therapy in SLE.

## Data Availability

Please contact Dr Vital to discuss data availability requests

## Acknowledgments and Affiliations

This research was supported by the United Kingdom National Institute for Health Research Leeds Biomedical Research Centre based at the Leeds Teaching Hospitals NHS Trust. The views expressed are those of the authors and not necessarily those of the NHS, the NIHR or the Department of Health. Dr Mahmoud was funded by the University of Benghazi. Dr Md Yusof is an NIHR Clinical Lecturer and Dr Vital is an NIHR Clinician Scientist. Dr. Ciurtin is funded by a Centre for Adolescent Rheumatology Versus Arthritis grant, 21593. There are no competing interests. This research was supported by a grant from LupusUK and the National Institute of Health Research (CS-2013-13-032).

## References

1. Mahmoud K, Zayat A, Vital EM. Musculoskeletal manifestations of systemic lupus erythmatosus. Current opinion in rheumatology. 2017; 29(5):486–492.

2. Pettersson S, Lovgren M, Eriksson LE, Moberg C, Svenungsson E, Gunnarsson I, et al. An exploration of patient-reported symptoms in systemic lupus erythematosus and the relationship to health-related quality of life. Scandinavian journal of rheumatology. 2012 Oct; 41(5):383–390.

3. Zayat AS, Md Yusof MY, Wakefield RJ, Conaghan PG, Emery P, Vital EM. The role of ultrasound in assessing musculoskeletal symptoms of systemic lupus erythematosus: a systematic literature review. Rheumatology. 2016; 55(3):485–494.

4. Isenberg DA, Rahman A, Allen E, Farewell V, Akil M, Bruce IN, et al. BILAG 2004. Development and initial validation of an updated version of the British Isles Lupus Assessment Group’s disease activity index for patients with systemic lupus erythematosus. Rheumatology (Oxford, England). 2005 Jul; 44(7):902–906.

5. Gladman DD, Ibanez D, Urowitz MB. Systemic lupus erythematosus disease activity index 2000. The Journal of rheumatology. 2002 Feb; 29(2):288–291.

6. Ibanez D, Gladman D, Urowitz M. Summarizing disease features over time: II. Variability measures of SLEDAI-2K. The Journal of rheumatology. 2007 Feb; 34(2):336–340.

7. NICE. Belimumab for treating active autoantibody-positive systemic lupus erythematosus. 2016 [cited; Available from:

8. Zayat AS, Md Yusof MY, Wakefield RJ, Conaghan PG, Emery P, Vital EM. The role of ultrasound in assessing musculoskeletal symptoms of systemic lupus erythematosus: a systematic literature review. Rheumatology (Oxford, England). 2016 Mar; 55(3):485–494.

9. Zayat AS, Mahmoud K, Md Yusof MY, Mukherjee S, D’agostino M-a, Hensor EM, et al. Defining inflammatory musculoskeletal manifestations in systemic lupus erythematosus. Rheumatology. 2018; 58(2):304–312.

10. Petri M, Orbai AM, Alarcon GS, Gordon C, Merrill JT, Fortin PR, et al. Derivation and validation of the Systemic Lupus International Collaborating Clinics classification criteria for systemic lupus erythematosus. Arthritis and rheumatism. 2012 Aug; 64(8):2677–2686.

11. McElhone K, Abbott J, Shelmerdine J, Bruce IN, Ahmad Y, Gordon C, et al. Development and validation of a disease-specific health-related quality of life measure, the LupusQoL, for adults with systemic lupus erythematosus. Arthritis Care & Research. 2007; 57(6):972–979.

12. D’Agostino M-A, Terslev L, Aegerter P, Backhaus M, Balint P, Bruyn GA, et al. Scoring ultrasound synovitis in rheumatoid arthritis: a EULAR-OMERACT ultrasound taskforce—Part 1: definition and development of a standardised, consensus-based scoring system. RMD open. 2017; 3(1):e000428.

13. Mahmoud K, Zayat AS, Yusof Y, Hensor E, Conaghan PG, Emery P, et al. Responsiveness of clinical and ultrasound outcome measures in musculoskeletal systemic lupus erythematosus. Rheumatology. 2019; 58(8):1353–1360.

14. van Tuyl LH, Lems WF, Boers M. Measurement of stiffness in patients with rheumatoid arthritis in low disease activity or remission: a systematic review. BMC musculoskeletal disorders. 2014 Jan 29; 15:28.

15. Vliet Vlieland TP, Zwinderman AH, Breedveld FC, Hazes JM. Measurement of morning stiffness in rheumatoid arthritis clinical trials. Journal of clinical epidemiology. 1997 Jul; 50(7):757–763.

16. Benlidayi IC. Fibromyalgia interferes with disease activity and biological therapy response in inflammatory rheumatic diseases. Rheumatology International. 2020:1–10.

17. Clark S, Tindall E, Bennett RM. A double blind crossover trial of prednisone versus placebo in the treatment of fibrositis. The Journal of rheumatology. 1985 Oct; 12(5):980–983.

18. Torrente-Segarra V, Salman-Monte T, Rúa-Figueroa I, Pérez-Vicente S, López-Longo F, Galindo-Izquierdo M. RELESSER Study Group of the Spanish Society of Rheumatology (SER) and the Study Group of Systemic Autoimmune Diseases of the SER (EAS-SER). Fibromyalgia prevalence and related factors in a large registry of patients with systemic lupus erythematosus. Clin Exp Rheumatol. 2016; 34(Suppl 96):S40–47.

19. Boers M, Kirwan JR, Wells G, Beaton D, Gossec L, d’Agostino MA, et al. Developing core outcome measurement sets for clinical trials: OMERACT filter 2.0. Journal of clinical epidemiology. 2014 Jul; 67(7):745–753.

20. Iagnocco A, Ceccarelli F, Rizzo C, Truglia S, Massaro L, Spinelli FR, et al. Ultrasound evaluation of hand, wrist and foot joint synovitis in systemic lupus erythematosus. Rheumatology (Oxford, England). 2014 Mar; 53(3):465–472.

21. Salliot C, Denis A, Dernis E, Andre V, Perdriger A, Albert JD, et al. Ultrasonography and detection of subclinical joints and tendons involvements in Systemic Lupus erythematosus (SLE) patients: A cross-sectional multicenter study. Joint, bone, spine: revue du rhumatisme. 2018 Dec; 85(6):741–745.

22. Murphy G, Isenberg DA. New therapies for systemic lupus erythematosus - past imperfect, future tense. Nat Rev Rheumatol. 2019 Jul; 15(7):403–412.

23. Wallace DJ, Stohl W, Furie RA, Lisse JR, McKay JD, Merrill JT, et al. A phase II, randomized, double-blind, placebo-controlled, dose-ranging study of belimumab in patients with active systemic lupus erythematosus. Arthritis and rheumatism. 2009 Sep 15; 61(9):1168–1178.

24. Navarra SV, Guzman RM, Gallacher AE, Hall S, Levy RA, Jimenez RE, et al. Efficacy and safety of belimumab in patients with active systemic lupus erythematosus: a randomised, placebocontrolled, phase 3 trial. Lancet (London, England). 2011 Feb 26; 377(9767):721–731.

25. Furie R, Petri M, Zamani O, Cervera R, Wallace DJ, Tegzova D, et al. A phase III, randomized, placebo-controlled study of belimumab, a monoclonal antibody that inhibits B lymphocyte stimulator, in patients with systemic lupus erythematosus. Arthritis and rheumatism. 2011 Dec; 63(12):3918–3930.

26. Aguiar R, Araujo C, Martins-Coelho G, Isenberg D. Use of Rituximab in Systemic Lupus Erythematosus: A Single Center Experience Over 14 Years. Arthritis Care Res (Hoboken). 2017 Feb; 69(2):257–262.

27. Merrill JT, Neuwelt CM, Wallace DJ, Shanahan JC, Latinis KM, Oates JC, et al. Efficacy and safety of rituximab in moderately-to-severely active systemic lupus erythematosus: the randomized, double-blind, phase II/III systemic lupus erythematosus evaluation of rituximab trial. Arthritis and rheumatism. 2010 Jan; 62(1):222–233.

28. Duxbury B, Combescure C, Chizzolini C. Rituximab in systemic lupus erythematosus: an updated systematic review and meta-analysis. Lupus. 2013 Dec; 22(14):1489–1503.

29. Md Yusof MY, Shaw D, El-Sherbiny YM, Dunn E, Rawstron AC, Emery P, et al. Predicting and managing primary and secondary non-response to rituximab using B-cell biomarkers in systemic lupus erythematosus. Ann Rheum Dis. 2017 Nov; 76(11):1829–1836.

30. Furie RA, Morand EF, Bruce IN, Manzi S, Kalunian KC, Vital EM, et al. Type I interferon inhibitor anifrolumab in active systemic lupus erythematosus (TULIP-1): a randomised, controlled, phase 3 trial. The Lancet Rheumatology. 2019; 1(4):e208–e219.

31. Morand EF, Furie R, Tanaka Y, Bruce IN, Askanase AD, Richez C, et al. Trial of Anifrolumab in active systemic lupus erythematosus. New England Journal of Medicine. 2019.

32. Ball EM, Tan AL, Fukuba E, McGonagle D, Grey A, Steiner G, et al. A study of erosive phenotypes in lupus arthritis using magnetic resonance imaging and anti-citrullinated protein antibody, anti-RA33 and RF autoantibody status. Rheumatology (Oxford, England). 2014 Oct; 53(10):1835–1843.

33. Coskun Benlidayi I. Fibromyalgia interferes with disease activity and biological therapy response in inflammatory rheumatic diseases. Rheumatology International. 2020 2020/06/01; 40(6):849–858.

